# Organ fat, not general obesity, defines risk for diabetes, inflammation, and comorbidities

**DOI:** 10.64898/2025.12.22.25342850

**Authors:** Matthew Ennis, Hajime Yamazaki, Shinichi Tauchi, Fumitaka Nakamura, Mitsuru Dohke, Nagisa Hanawa, Robert Wagner, Martin Heni

## Abstract

**Background:** Fat distribution patterns, rather than total adiposity alone, critically influence the risk of obesity-related diseases. However, due to correlations between fat accumulation in different regions, the contribution of regional fat to metabolic and non-metabolic diseases remains unclear.

**Methods:** Using UK Biobank MRI data (N = 23,548) and a Japanese cohort (N = 642), we used archetype analysis to identify patterns of predominantly isolated fat accumulation in the liver, pancreas, visceral adipose tissue, and thigh muscle. We then characterized the type 2 diabetes (T2D) risk, biomarker profiles, and broad health burden associated with isolated fat accumulation.

**Findings:** We identified four distinct patterns of isolated fat accumulation in the liver, thigh muscle, pancreas, and visceral adipose tissue and replicated these patterns in the Japanese cohort. Organ-specific fat accumulation was associated with an equal or greater T2D burden than isolated visceral adiposity, despite lower BMI and visceral fat amount. Moreover, specific fat depots were linked to distinct comorbidities not observed with visceral fat alone, including knee osteoarthritis (thigh myosteatosis), COPD (pancreatic steatosis) and breast cancer (hepatic steatosis). In contrast, total and visceral fat alone were not significantly associated with many of these complications.

**Interpretation:** These findings highlight the central role of organ-specific fat accumulation beyond general adiposity in obesity-related diseases, offering new insight into the heterogeneity of obesity.

**Funding:** European Research Council, European Union, and the Japanese Society for the Promotion of Science.

## Introduction

Obesity is a leading driver of morbidity^1^ and mortality^2^, though the health risks it confers are not solely determined by total fat mass^3^. Instead, the distribution of fat, particularly in visceral adipose tissue (VAT)^4^, the liver^5^, the pancreas^6^, and skeletal muscles^7^, have been linked to increased risk of type 2 diabetes (T2D)^8^. However, because fat levels across these regions are often correlated, it remains challenging to disentangle their individual contributions to metabolic and non-metabolic diseases.

The UK Biobank (UKB) Imaging Project has collected magnetic resonance imaging (MRI) data for up to 100,000 participants^9^, providing an unparalleled opportunity to analyze whole-body fat distribution and its effect on human health. To characterize patterns of fat distribution in the UKB cohort, we applied archetype analysis^10^. This technique identifies extreme patterns in multivariate data. In a previous study, we used unsupervised clustering to identify fat distribution patterns in two small cohorts, demonstrating that distinct distribution profiles confer markedly different risks for T2D^8^. Yet, these clusters reflected patterns of co-accumulation of fat across multiple depots and were not designed to isolate the effects of fat accumulation within a single depot.

In this study we use archetype analysis to disentangle the health effects of fat accumulation across multiple organs, enabling a mechanistic view of fat distribution and disease. Specifically, we used archetype analysis to identify groups of UKB participants with isolated fat accumulation within specific depots and investigated the health impacts associated with each distribution pattern. To assess generalizability, we further evaluated the presence of these fat distribution patterns in an independent Japanese cohort. Together, this approach provides a comprehensive assessment of how organ-specific fat accumulation, rather than general obesity alone, shapes disease risk.

## Results

### Archetypes of body fat distribution

We performed archetype clustering on MRI-quantified and sex-scaled fat measurements from thigh muscles, visceral adipose tissue, pancreas, and liver in 23,548 UK Biobank participants. This analysis revealed five archetypes, each representing an extreme of specific body fat distribution: thigh myosteatosis, visceral adiposity, pancreatic steatosis, and hepatic steatosis, as well as a phenotype with low fat accumulation across all depots (Steatopenia) (Figure 1A; Supplementary figure 1). The same extremes were identified when clustering was performed without sex-stratified scaling (Supplementary figure 2).

**Figure 1.**
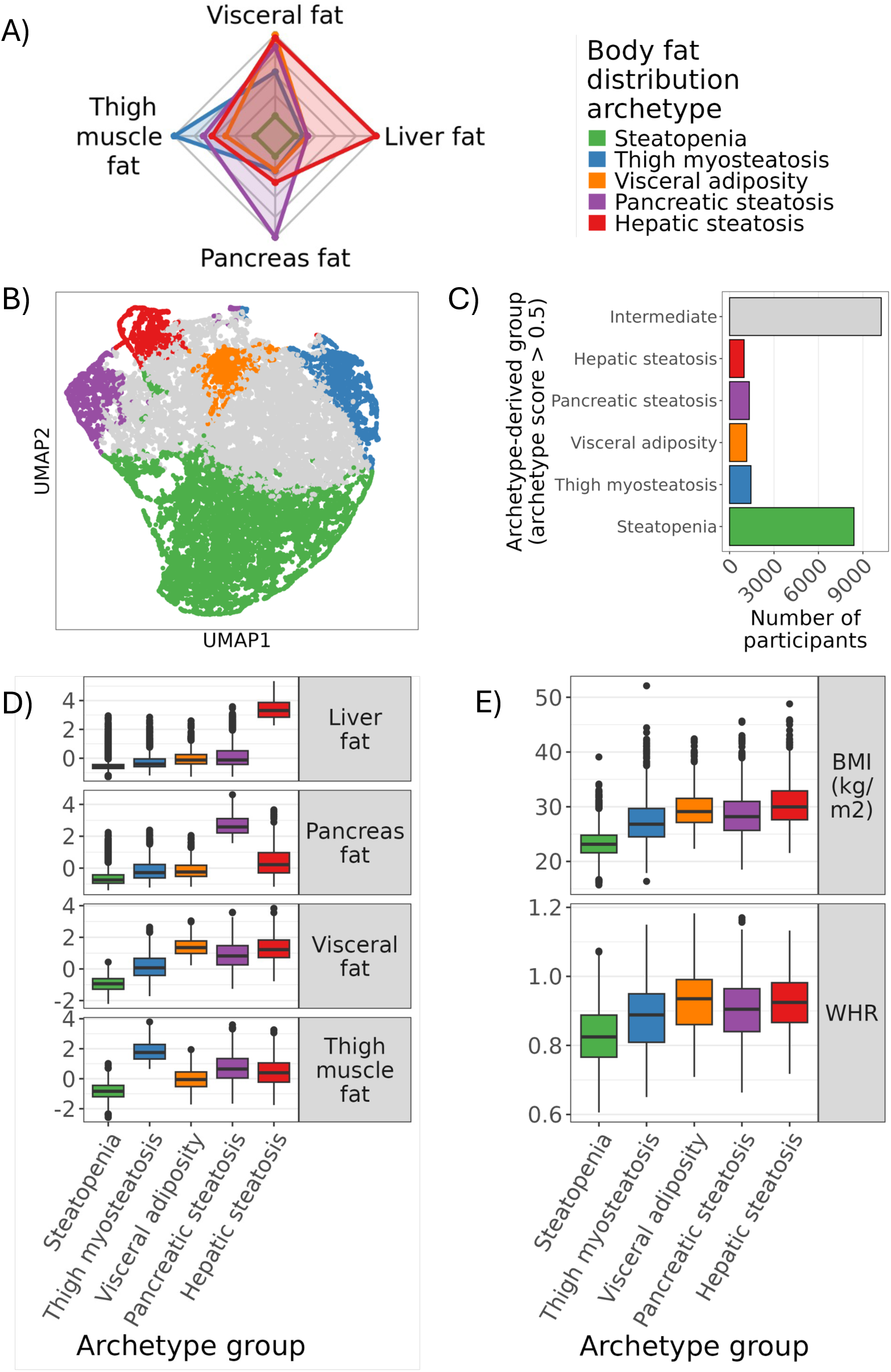
Archetype clustering of the UK Biobank MRI cohort identifies individuals with extreme body fat distributions. Using 23,548 participants from the UK Biobank MRI cohort, we performed archetype clustering on MRI-quantified levels of fat accumulation in the liver, pancreas, thigh muscle, and visceral adipose tissue. **(A)** Identified archetypes of body fat distribution. **(B)** UMAP projection of individuals who are most similar to each identified archetype (‘archetype groups’). **(C)** Number of individuals most similar to each archetype or who are intermediate in similarity between archetypes. **(D)** Distributions of fat levels in each depot across archetype groups. **(E)** Distributions of body mass index (BMI) and waist-to-hip ratio (WHR) across archetype groups.

Each participant received five archetype scores ranging from 0 to 1, reflecting their similarity to each archetype. These scores summed to 1 for each individual, enabling the identification of those with predominantly isolated fat accumulation (defined as a single archetype score > 0.5, indicating dominant similarity to one extreme pattern) (Figure 1B). This approach identified 1,444 (6.13%), 1,160 (4.93%), 1,331 (5.65%), and 981 (4.17%) individuals within *Thigh myosteatosis*, *Visceral adiposity*, *Pancreatic steatosis*, and *Hepatic steatosis* archetype groups, respectively, while 8390 (35.63%) individuals exhibited low fat content in these 4 depots (*Steatopenia*). The remaining 10,242 (43.49%) showed intermediate fat distribution profiles (‘Intermediate’) (Figure 1C and 1D; Supplementary tables 1 and 2).

Body mass index (BMI) and waist-to-hip ratio (WHR) were elevated in all archetype groups relative to the Steatopenia group (adj.p < 0.005 for all comparisons), but the degree of elevation varied, underscoring the limited ability of these metrics to reflect depot-specific fat accumulation (Figure 1E and Supplementary table 3). Strikingly, 30% and 20% of individuals within the Thigh myosteatosis and Pancreatic steatosis groups had a ‘healthy’ BMI less than 25kg/m^2^ (Supplementary table 4 and Supplementary figure 3).

In summary, we identified four archetypes with extreme fat accumulation in the thigh muscles, visceral adipose tissue, pancreas, or liver, and a fifth group with minimal fat in these depots.

### Archetype replication in Japanese cohort

To validate our findings, we replicated the occurrence of extreme forms of localized fat accumulation in an independent cohort of Japanese patients (N = 642) (Supplementary figure 4 and Supplementary table 5). As in the UK Biobank cohort, individuals in the Japanese Trunk myosteatosis, Pancreatic steatosis, and Hepatic steatosis groups, showed pronounced and localized fat accumulation in muscle, the pancreas, and the liver (Supplementary Figure 4D), respectively, but at lower BMI levels (Supplementary figure 4E).

### T2D burden of individuals with extreme body fat distributions

We next tested associations of extreme body fat distributions with prevalent (Figure 2A) and incident T2D (Figure 2B), using the Steatopenia group as reference.

**Figure 2.**
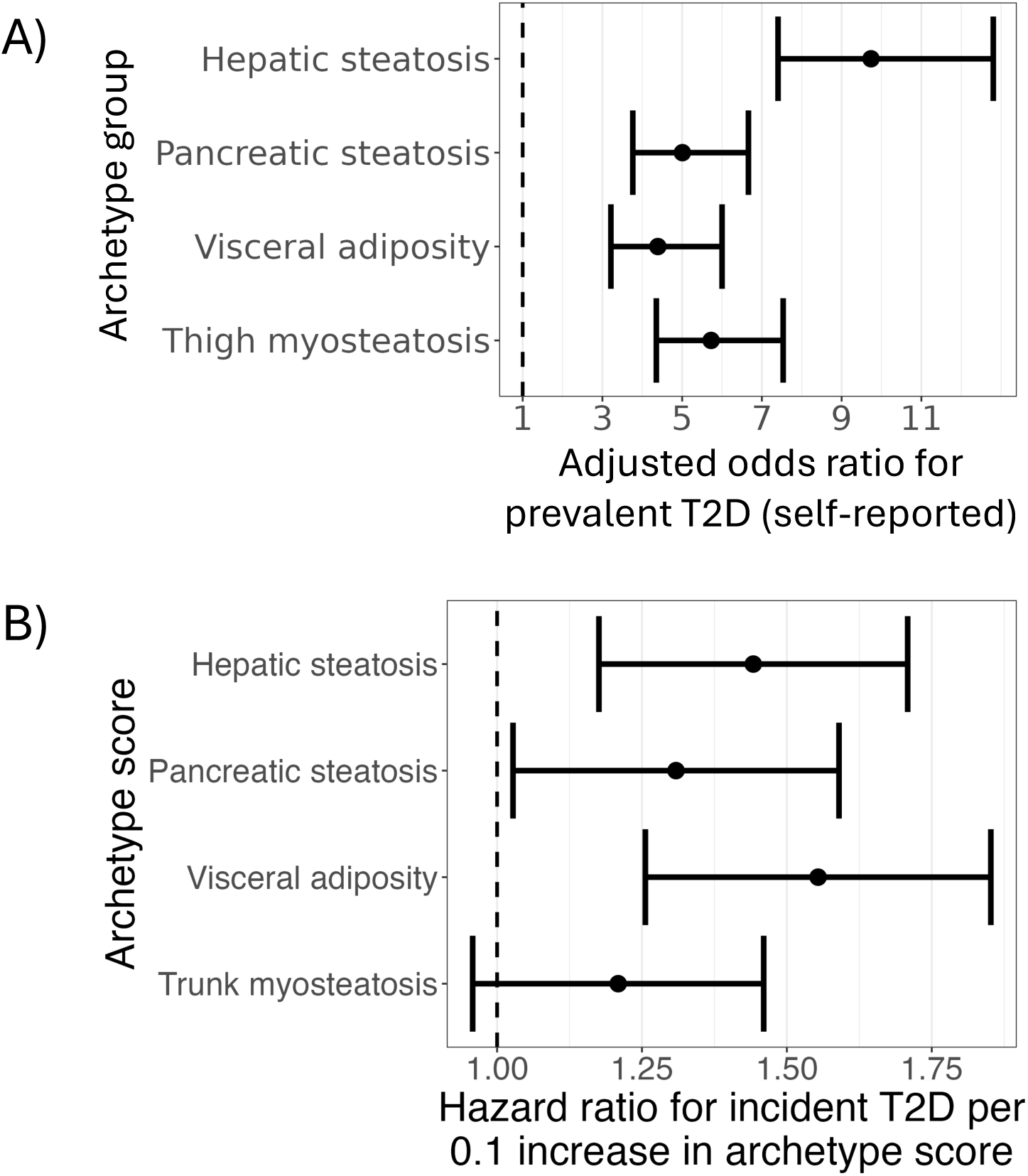
Type 2 diabetes (T2D) burden of extreme forms of fat accumulation. We compared the associations of the identified archetype groups characterised by extreme forms of body fat accumulation with prevalent **(A)** and incident (**B**) T2D. (**A**) Associations of extreme fat accumulation archetype groups with prevalent T2D in the UK Biobank cohort (N = 23,548). Effects were calculated relative to the Steatopenia group and were adjusted for age, sex, smoking status, and alcohol intake frequency. (**B**) Associations of extreme fat accumulation archetype scores with incident T2D in the Japanese replication cohort (N = 642). Whiskers represent 95% confidence intervals.

There were 1,057 prevalent cases of T2D across the UK Biobank cohort, with 398 (37.65%) of these cases within one of the 4 archetype groups. Each archetype group was significantly associated with prevalent T2D, with the Hepatic steatosis group showing the largest effect (OR = 9.74, adj.p < 0.005) (Figure 2A). Strikingly, despite substantially lower BMI, WHR, and visceral fat levels, both the Pancreatic steatosis and Thigh myosteatosis groups were associated with prevalent T2D at levels comparable to the Visceral adiposity group (adj.p > 0.05 for all pairwise comparisons) (Supplementary table 6). Extreme fat accumulation archetype scores in the Japanese cohort were significantly associated with an increased risk of incident T2D (N = 49), including Hepatic steatosis, Visceral adiposity, and Pancreatic steatosis (p < 0.05 for all) (Figure 2B). Trunk myosteatosis had a trend towards an increased incident T2D (p = 0.07).

These results show that individuals with significant fat accumulation in the pancreas, liver, or muscle face elevated T2D burden, even at a lower BMI, WHR, and visceral fat levels than those with predominantly only visceral fat accumulation.

### Clinical biomarker and immune cell profiles of extreme fat accumulation before T2D onset

To assess the early clinical impact of extreme forms of fat accumulation, we examined biomarkers (Figure 3A; Supplementary table 7) and immune cell profiles (Figure 3B; Supplementary table 8) in individuals without pre-existing T2D.

**Figure 3.**
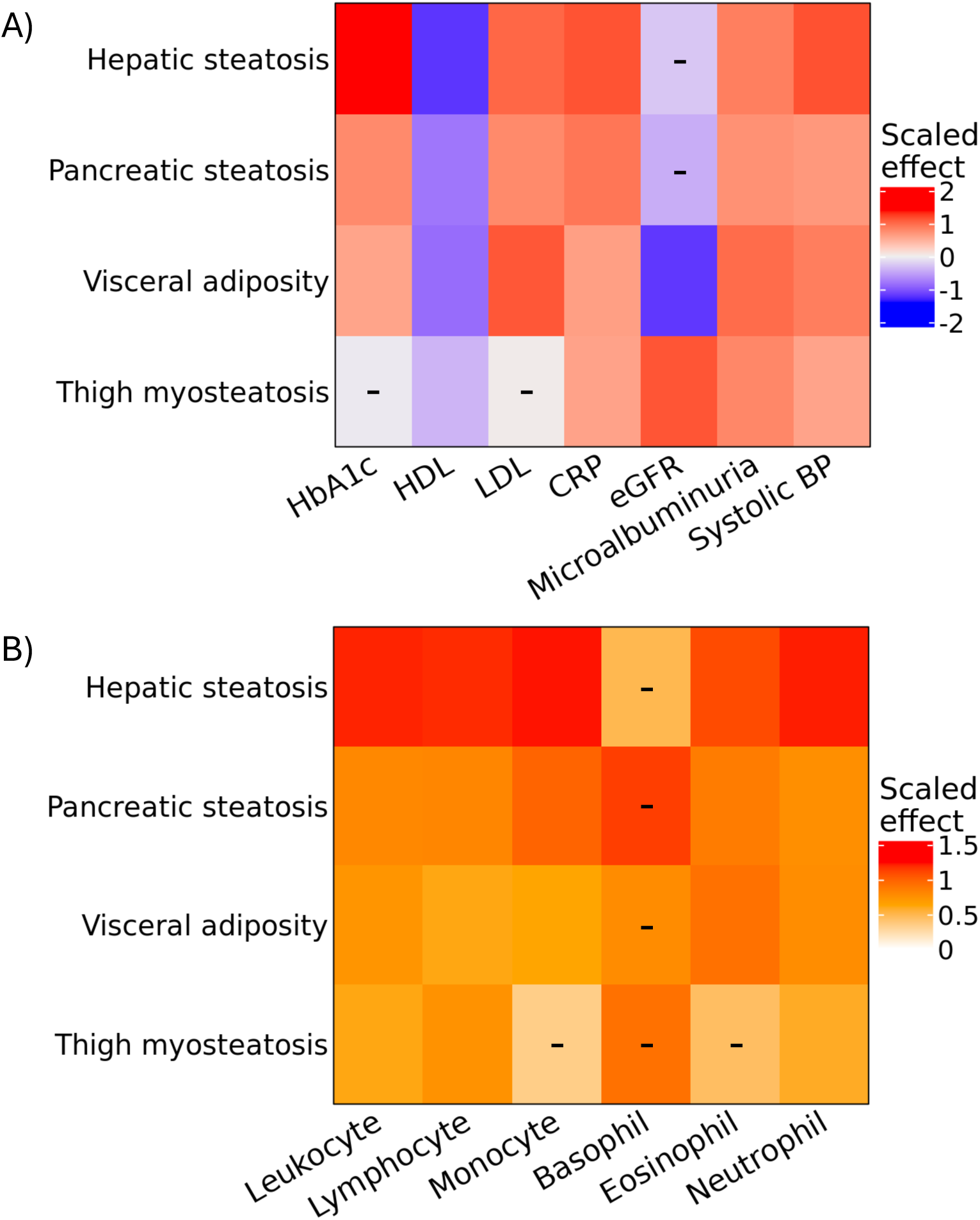
Clinical biomarker presentation of pre-T2D extreme body fat distribution groups. Associations of archetype groups with biomarker levels in individuals from the UK Biobank without prevelent T2D allowed the comparison of the contribution of fat accumulation in each depot to clinical presentation. Effects were calculated relative to the Steatopenia group and were adjusted for age, sex, smoking status, alcohol intake frequency, and time elapsed between initial and imaging UKB visits. Dashed lines indicate a non-significant difference compared to the Steatopenia group. **(A)** Adjusted associations of archetype groups with clinical biomarkers in individuals without prevalent T2D. Effects on HDL and LDL levels were estimated in individuals not currently on cholesterol-lowering medication. Effects on CRP were estimated in individuals with a CRP level < 5 mg/L. Effects on systolic blood pressure were further adjusted by participant current antihypertensive medication usage status (yes/no). **(B)** Adjusted associations of archetype groups with immune cell counts in individuals without prevalent T2D. A dash (‘-’) within the heatmap indicates the association was non-significant (adj.p > 0.05).

Biomarker profiles differed substantially across archetype groups (Figure 3A). The Hepatic steatosis exhibited the most unfavourable profile. CRP was markedly higher in both the Pancreatic steatosis (1.36 mg/L increase, adj.p < 0.005) and Hepatic steatosis (1.69mg/L increase, adj.p < 0.005) groups than with the Visceral adiposity group (0.93mg/L, adj.p < 0.005). Furthermore, CRP elevation in the Thigh myosteatosis group (0.89mg/L increase, adj.p < 0.005) was comparable to the Visceral adiposity group, further indicating a substantial contribution of organ-specific fat to systemic inflammation (Figure 3A; Supplementary table 7).

Increased LDL and decreased HDL cholesterol were observed in all groups except Thigh myosteatosis, which exhibited only a more minor reduction in HDL. HDL cholesterol was markedly lower in Hepatic steatosis than other groups (Figure 3A).

Counts of leukocytes, lymphocytes, and neutrophils were significantly elevated in all groups (adj.p < 0.001), while counts of monocytes and eosinophils were elevated in all groups (adj.p < 0.005) but the Thigh myosteatosis group (Figure 3B; Supplementary table 8). The Hepatic steatosis group presented with the most severe immune cell profile, which exhibited the largest elevation in all cell counts except basophils (adj.p > 0.05). The Pancreatic steatosis group also showed a greater elevation in monocytes compared to the Visceral adiposity group, further emphasizing the pro-inflammatory role of organ-specific fat (Figure 3B).

### Prevalent comorbid associations of extreme body fat distributions

To investigate the broader health impacts of fat distribution beyond type 2 diabetes, we assessed associations between the four archetype groups and the prevalence of ICD10-coded diagnoses (Figure 4; Supplementary tables 9 and 10).

**Figure 4.**
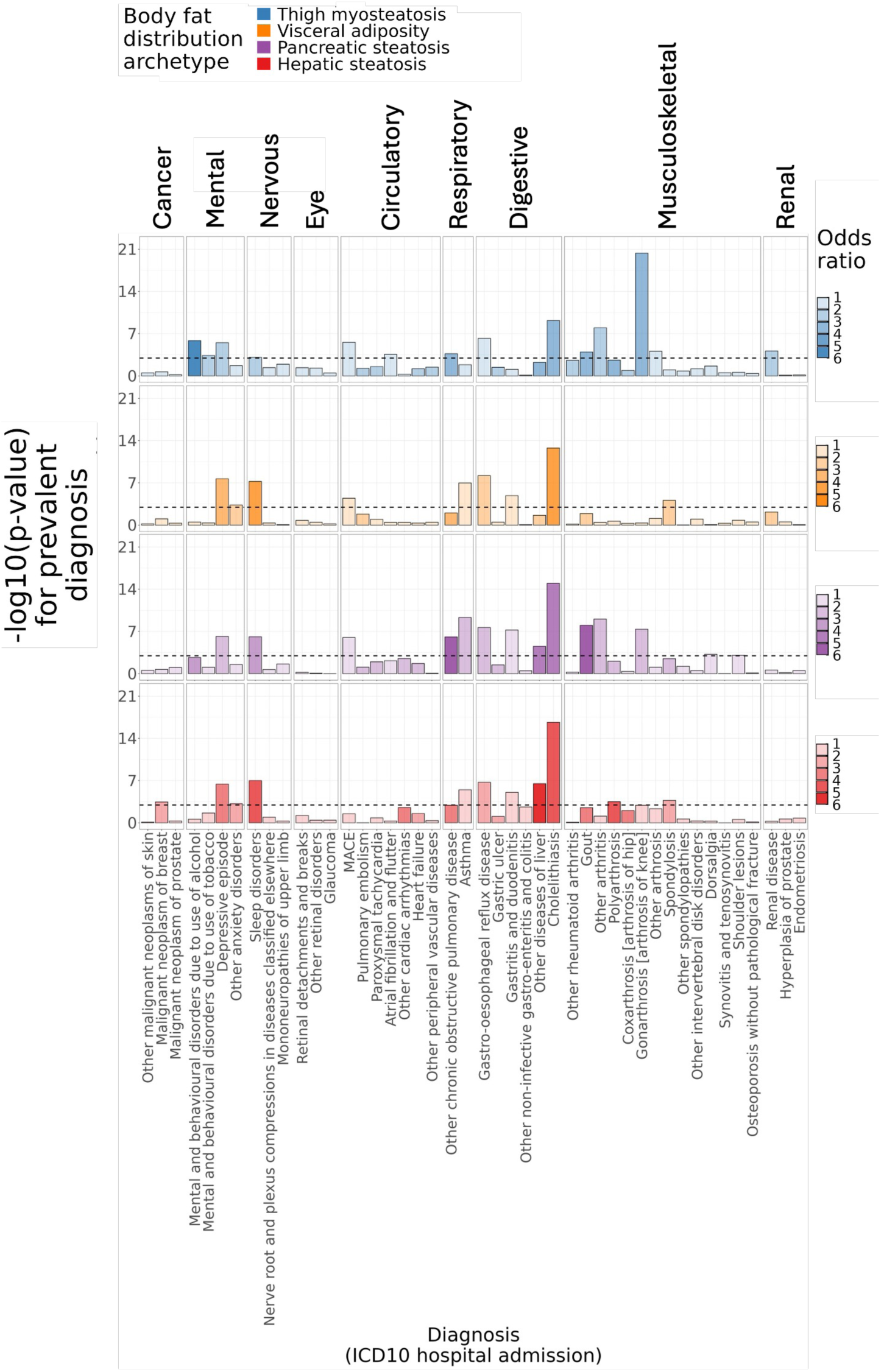
Screening of prevalent diagnoses identifies distinct comorbidities observed with organ-specific fat accumulation. We compared the associations of archetype groups with prevalent (>1% of participants) ICD10 diagnoses derived from UK Biobank hospital inpatient records. Effects were calculated relative to the Steatopenia group and were adjusted for age, sex, smoking status, and alcohol intake frequency. Plotted are bar charts showing the p-value of the association of each archetype group with each respective diagnosis. The dashed line indicates that Bonferroni-adjusted significance levels for the number of diagnoses tested (45). Bar charts are colored by the the respective archetype group, The strength of color is varied by the adjusted odds ratio of each respective group-diagnosis association. ICD10 diagnoses are grouped by their respective ICD10 chapter representing the affected high-level physiological system.

While all archetype groups showed elevated rates of depression, sleep disorders, reflux, and gallstone disease (p < 0.001), each was characterize**d** a distinct pattern of comorbidities.

The Thigh myosteatosis group exhibited a significant burden of multiple diseases, with markedly increased prevalence of MACE (OR = 1.79, p < 0.001, N = 547), renal disease (OR = 2.55, p < 0.001, N = 64), and knee osteoarthritis (OR = 3.82, p < 0.001, N = 143). This group also had higher rates of COPD, atrial fibrillation, and mental disorders related to alcohol and tobacco use (p < 0.001 for all) (Figure 4; Supplementary tables 9 and 10).

The Visceral adiposity group likewise showed an increased burden of MACE (OR = 1.84, p < 0.001, N = 275) and additionally a greater prevalence of anxiety, asthma, and spondylosis (p < 0.001 for all). However, unlike Thigh myosteatosis, Visceral adiposity was not associated with arthritis or joint disease, suggesting a preserved musculoskeletal health.

The Pancreatic steatosis group showed a distinct pattern, with strong associations with chronic respiratory diseases, including COPD (OR = 5.47, p < 0.001, N = 78) and asthma (OR = 2.14, p < 0.001, N = 299). This group also showed increased prevalence of liver disease (OR = 4.56, p < 0.001), as well as MACE, gout, arthritis, and knee osteoarthritis (p < 0.001 for all).

As with Visceral adiposity, the Hepatic steatosis group had elevated rates of anxiety, asthma, and spondylosis. In addition, this group was strongly (OR = 4.09) associated with polyarthrosis (p < 0.001, N = 13), consistent with the marked systemic inflammation observed in this group (Figure 3). Hepatic steatosis also uniquely exhibited elevated breast cancer prevalence (OR = 2.16, p < 0.001, N = 94) (Figure 4; Supplementary tables 9 and 10). Interestingly, despite having a BMI and visceral fat mass similar to the Visceral adiposity group, the Hepatic steatosis group was not significantly associated with MACE, suggesting a potentially different cardiovascular risk profile (Figure 4; Supplementary tables 9 and 10).

Overall, these findings reveal that organ-specific fat is a primary determinant of complication burden, highlighting the importance of fat distribution, rather than total adiposity alone, in shaping chronic disease profiles in obesity.

## Discussion

We identified four distinct patterns of body fat distribution characterized by pronounced fat accumulation in specific depots, i.e. visceral adipose tissue, pancreas, liver, and thigh muscles. Each pattern was associated with a unique clinical and metabolic profile. These findings emphasize that obesity is not a uniform condition^11^ and that the health consequences of excess fat depend critically on where fat is stored.

Our results highlight that organ-specific fat depots frequently carry greater metabolic risk than visceral fat. Although all four archetypes were associated with increased type 2 diabetes (T2D), the magnitude of risk and the phenotypic presentation varied markedly by depot. Notably, individuals with extreme thigh myosteatosis had T2D burden equivalent to those with extreme visceral fat, despite having a lower BMI and substantially less visceral fat. These findings underscore the limited ability of conventional anthropometric indices to capture metabolically adverse fat distributions and highlight the potential clinical value of assessing fat distribution to refine risk stratification.

An unexpected finding was the comparatively milder inflammatory profile observed with extreme visceral fat accumulation, especially in comparison to hepatic and pancreatic steatosis. This challenges the long-standing view that visceral fat is the primary driver of systemic inflammation^12^. Instead, our findings suggests that visceral fat alone may not be sufficient to trigger a prominent inflammatory response in the absence of concurrent fat in other organs, particularly the liver.

Beyond T2D, organ-specific fat accumulation was associated with a distinct burden of several comorbidities that were not seen in isolated visceral adiposity. In particular, arthrosis/arthritis of any kind was not significantly elevated in the Visceral adiposity group but was elevated in groups with organ-specific fat, especially thigh myosteatosis. This finding emphasizes that body fat distribution, and not just total adiposity, is a significant determinant of musculoskeletal health risk^13,14^.

Furthermore, pancreatic steatosis was strongly associated with COPD, independent of smoking. Although the mechanisms linking pancreatic fat to pulmonary disease remain unclear, but systemic inflammation might be an important link^15^. Interestingly, pancreatic steatosis, and not hepatic steatosis, has also been linked to the severity of COVID-19 pneumonia, suggesting a specific link between pancreatic fat and lung vulnerability^16^.

In addition to its metabolic and inflammatory effects, hepatic steatosis was associated with increased breast cancer prevalence. This is in line with experimental evidence in rodents demonstrating that hepatic steatosis results in the upregulation of several hepatokines, including FGF21, which promote breast cancer tumor growth^17^. Our results indicate that similar endocrine effects of liver fat might be present in humans.

Surprisingly and in contrast to other archetypes, the Hepatic steatosis group was not significantly associated with prevalent MACE after adjustment for confounders. This observation is consistent with a recent UK Biobank study showing that liver impairment (assessed through increased iron-corrected T1 relaxation time) was a robust predictor of cardiovascular risk, whereas liver fat itself was not^18^. Furthermore, genetic evidence supports the hypothesis that liver fat may, under certain conditions, even be protective against cardiovascular disease^19^. One potential mechanism involves the upregulation of the hepatokine FGF21 in hepatic steatosis, which was shown to protect from atherosclerosis^20^ and vascular impairment^21^. Together these results suggest that liver fat accumulation on average may be protective of cardiovascular disease. However, this hypothesis remains speculative, and further work is needed to clarify the causal role of liver fat in cardiovascular outcomes.

There are limitations to our work. Despite replicating these patterns of extreme fat accumulation in a small independent cohort of Japanese patients, the predominantly European ethnic background of the UKB cohort potentially limits generalizability of the associated health impacts of these patterns. Additionally, we limited our analysis to prevalent hospital diagnoses as the relatively short post-imaging visit follow-up available precluded a broad view of incident outcomes. Furthermore, biomarkers and immune cell counts were collected several years before imaging, and our adjustments for the time difference between visits may not fully control for this.

In conclusion, we identified four distinct patterns of extreme body fat accumulation in the liver, pancreas, thigh muscle, and visceral depot. Organ-specific fat accumulation was associated with an equal or greater burden of T2D, greater systemic inflammation, and a higher burden of comorbidities than isolated visceral adiposity, despite frequently lower BMI and less visceral fat. These findings highlight the clinical relevance of body fat distribution and underscore the health risks associated with organ-specific fat, which are not adequately captured by general measures of adiposity.

## Methods

### UKB MRI study

The UK Biobank (UKB) Imaging Project is a cohort study aiming to acquire whole-body magnetic resonance imaging (MRI) images of 100,000 participants. Whole-body MRI enables the simultaneous quantification of fat accumulation across multiple depots.

### Depot fat quantification

Quantification of fat accumulation levels in the visceral adipose tissue (VAT), liver, pancreas, and thigh muscles have been previously described^22,23^. We accessed the following UKB variables: ‘Visceral fat volume (f.21085)’, ‘Liver PDFF (fat fraction) (f.21088)’ and ‘Pancreas PDFF (fat fraction) (f.21090)’. Furthermore, levels of thigh muscle fat infiltration were derived from taking the mean values of the 4 variables indicating anterior/posterior right/left thigh muscle fat infiltration (f.24353, f.24354, f.23355, f.23346).

### Exclusion criteria

Participants with missing values for any of the 4 fat depot amounts at the initial imaging assessment visit were excluded. Participants with pre-existing type 1 diabetes (T1D) or gestational diabetes by the time of the initial imaging visit were excluded. Multivariate outliers (N = 1321) of depot fat levels were removed using the Mahalanobis distance calculated within each sex. The threshold Mahalanobis distance to be considered an outlier was calculated as the quantile corresponding to a 1% probability of selecting a larger value from a chi-squared distribution with 4 degrees of freedom (number of fat depots considered). This resulted in a total cohort of 23,548 participants for this study.

### Archetype clustering

Depot fat variables were scaled by sex by dividing values by the depot fat standard deviation within each sex.

To determine the optimal number of archetypes, we first split the cohort based on the date of each participant’s imaging visit to ensure stability of archetypes to methodological changes which may have occurred over time. Archetype clustering was performed on the discovery and validation subsets separately. An elbow plot of the marginal variance explained for each archetype solution was plotted in discovery and validation to identify the optimal number of archetypes. Archetype analysis without sex-stratified scaling yielded similar archetypes but differences in archetype scores between men and women necessitated the use of sex scaling.

For each of the five identified archetypes, a score from 0 to 1 was derived for each participant indicating the similarity of their body fat distribution to each respective archetype (i.e. having extreme fat accumulation in that depot). Each participant’s five scores in total sum to 1. As an example, an individual with a single archetype score of 1 will have scores of 0 for the remaining archetype scores, indicating an individual with a body fat distribution maximally similar to a single extreme. In contrast, an individual with an archetype score of 0.2 for each of the 5 archetype scores will have a body fat distribution which is exactly intermediate of the 5 extremes.

To identify groups of participants characterized by extreme forms of fat accumulation, we selected for individuals with a score >0.5 for each of the five archetype scores, resulting in five separate groups. Participants with no archetype score >0.5 were assigned to an ‘Intermediate’ group.

Archetype analysis was performed using the ‘archetypal’ R package (version 1.3.1).

### Participant characteristics

Demographic and lifestyle data, including age, sex, BMI, waist circumference, hip circumference, smoking status, and alcohol intake frequency were collected at both the initial UKB assessment and the initial imaging center visit. Smoking status was reported as ‘Current’, ‘Previous’ and ‘Never’ while alcohol intake frequency was reported as ‘Daily or almost daily’, ‘Three or four times a week’, ‘Once or twice a month’, ‘Special occasions only’, and ‘Never’.

### Japanese CT study

The Japanese cohort is derived from a computed tomography (CT) study which included CT scans obtained from health examinations conducted between 2008 and 2013 at Keijinkai Maruyama Clinic in Sapporo, Japan. From 2,168 individuals without diabetes described previously^8^, 642 randomly selected individuals were analyzed. As in the UKB MRI study, archetype clustering was performed, and baseline characteristics of the participants were compared between clusters using Fisher’s exact test and Kruskal-Wallis test.

### Associations of archetype groups with type 2 diabetes

Prevalent T2D in the UKB MRI study was defined using self-reported questionnaires (f.2443). Individuals with ambiguous answers (not ‘Yes’ or ‘No’) were removed. Incident T2D in the Japanese CT study was defined as having at least one of: fasting plasma glucose reaching 126 mg/dL or higher, an HbA1c level of at least 6.5%, or use of antidiabetic medication. The hazard ratios for incident T2D were estimated using Cox proportional hazards models.

### Biomarkers/immune cell counts

All biomarker measurements and immune cell counts were taken at the initial UKB assessment center visit. To characterize pre-disease phenotypes, analyses were restricted to individuals without a diagnosis of diabetes (type 1 or 2) by the time of the imaging visit.

### Prevalent comorbid diagnoses

Prevalent ICD10 diagnoses were extracted from each participant’s hospital inpatient records and aggregated into 3-letter codes. We retained only diagnoses with >1% prevalence in the cohort. This list was further curated with clinician guidance (by authors MH and RW). Calculated p-values for the associations of each archetype group with prevalent diagnoses were Bonferroni-adjusted for the N = 45 of tested ICD10 diagnoses, resulting in a threshold of p < 0.001 (0.05/45) to be deemed significant. A compositive indicator for MACE was created by aggregating the Ischaemic heart diseases (I20, I21, and I25) and stroke (I63 and I67) codes. Acute and chronic renal failure (N17 and N18) were further aggregated into ‘Renal disease’.

### Statistical analyses

All statistical analyses in the UKB study were conducted using the statistical programming language R (version 4.4.0). The Steatopenia group was used as the reference group for all models, except for pairwise univariate tests directly comparing differences between archetype groups.

Group differences in continuous variables were assessed using two-tailed Wilcoxon signed-rank tests and categorical variables using two-tailed Chi-square tests.

Multivariate linear and logistic models were used to assess associations of archetype groups with continuous and categorical variables, respectively. All associations were adjusted for age, sex, smoking status, and alcohol intake. Associations for biomarker measurements and immune cell counts were adjusted using covariates measured at the initial UKB assessment center visit, while all other associations used covariates measured at the initial UKB imaging center visit. Analyses of HDL and LDL excluded individuals on lipid-lowering medications while C-reactive protein (CRP) analyses excluded individuals with CRP > 5 mg/L. Systolic blood pressure analyses were adjusted for current antihypertensive medication use (yes/no). Associations with biomarkers and immune cell counts were adjusted by the time elapsed between the participant’s initial assessment and their imaging visit (median = 9.54 years, IQR = 6.17-10.33 years).

All p-values were adjusted using Benjamini-Hochberg, except prevalent comorbidity associations which were adjusted using Bonferroni.

## Data availability and analysis scripts

UKB data is available only to approved researchers for which an application can be made at https://www.ukbiobank.ac.uk/use-our-data/apply-for-access/. The analysis script for reproducing the results and figures from UKB input data are available at https://github.com/MattEnnis74/UKB-fat-distribution-archetypes.

## Acknowledgements

This research has been conducted using the UK Biobank under application number 383378. We would like to thank all the participants of UK Biobank for their vital contribution to this data resource.

## Author Contributions

ME analyzed the data, created visualizations, drafted the manuscript text, and contributed to project conceptualization and discussions. HY and RW supported data analysis and contributed to project conceptualization and discussions. S.T., F.N., M.D., and N.H. collected data.MH supervised the project, contributed to manuscript drafting, project conceptualization and discussions. All authors reviewed, made critical revisions, and approved the final version of the manuscript prior to submission.

