## Supplementary figures for "Organ fat, not general obesity, defines risk for diabetes, inflammation, and comorbidities"

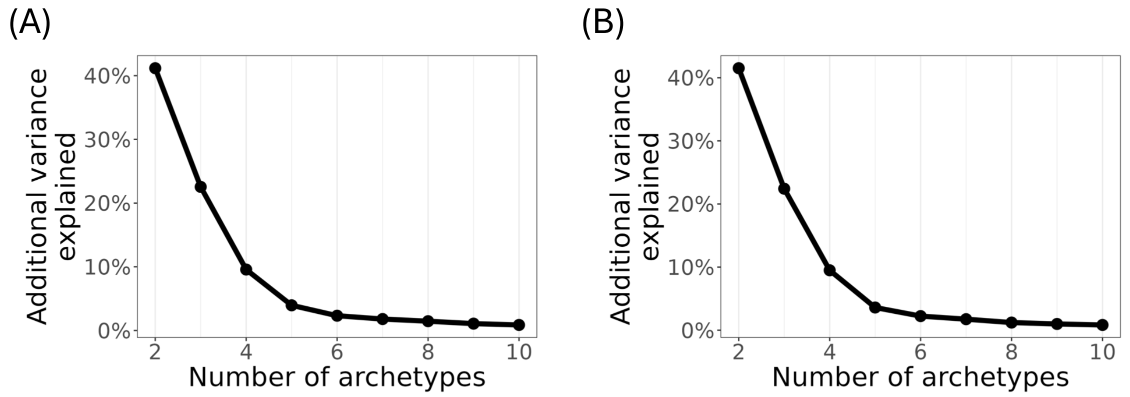


**Supplementary figure 1. Discovery and validation of archetypes.** Individuals were split equally according to the period of imaging center visit (2016-2018 and 2018-2020) into a discovery (A) (N = 12,421) and validation set (B) (N = 12,448), respectively. Archetype clustering was performed independently in each set to identify the optimal number number of archetypes to represent the whole cohort.

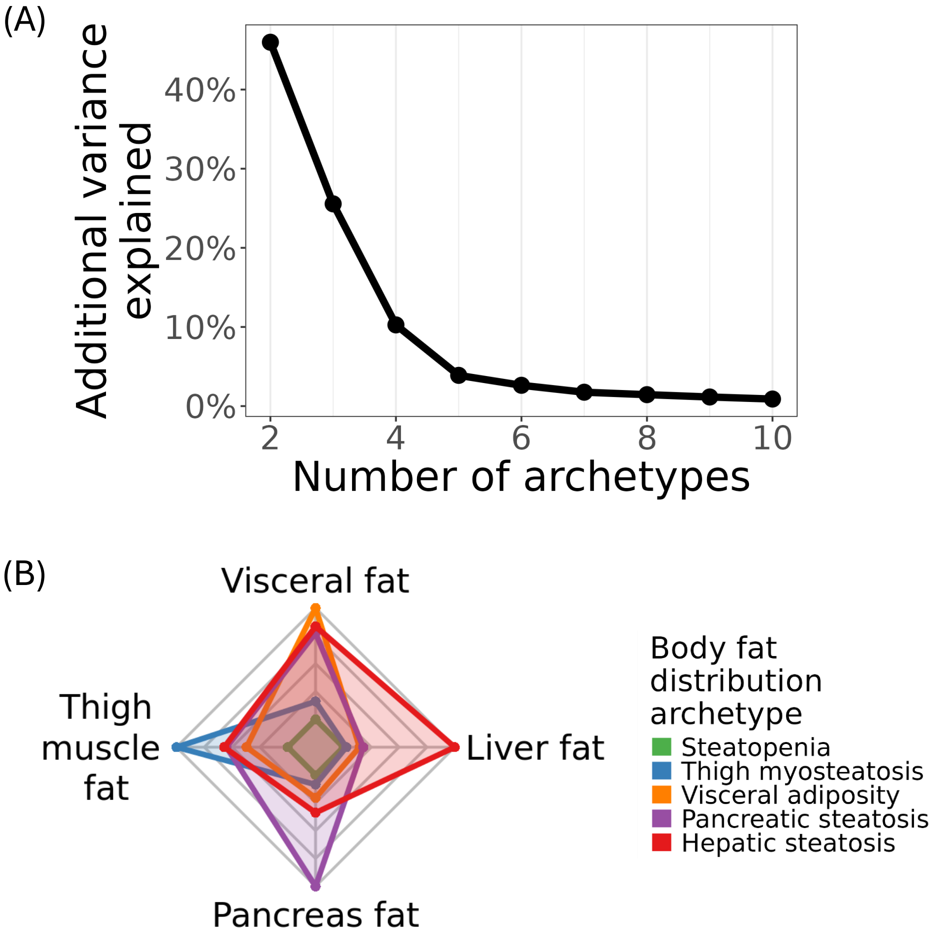


**Supplementary figure 2. Sensitivity analysis of archetype clustering without sex-stratified scaling.** We performed archetype clustering without sex-stratified scaling of fat depot levels to evaluate the effects on the final clustering solution. (A) Variance explained by archetype solutions for increasing numbers of archetypes. (B) Fat depot levels of the identified optimal number of archetypes (k = 5).

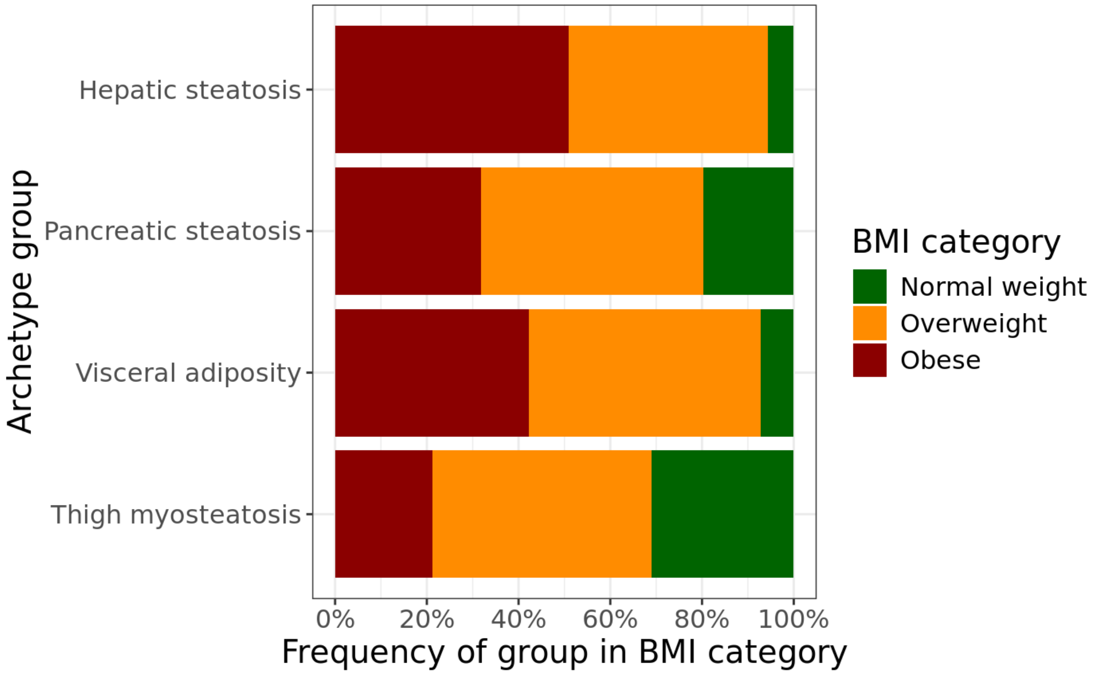


**Supplementary figure 3. Frequency of BMI categories in extreme body fat distribution groups.** Individuals within archetype groups were categorised into 3 BMI categories: normal weight (< 25kg/m$^2$), overweight (>= 25kg/m$^2$ and < 30kg/m$^2$), and obese (>= 30kg/m$^2$).

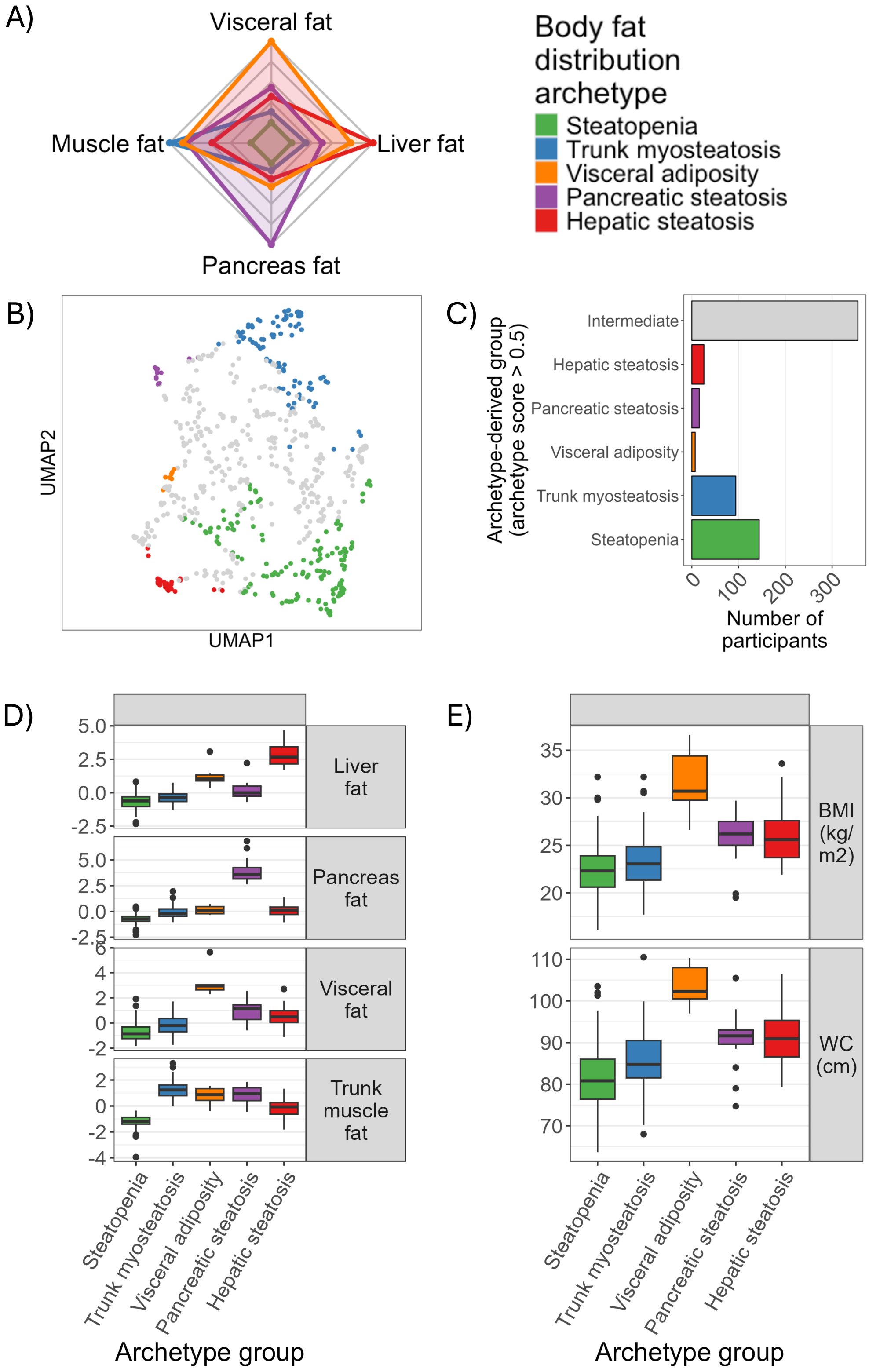


**Supplementary figure 4. Replication of archetypes in Japanese cohort.** We replicated the occurrence of extreme forms of localized fat accumulation in an independent cohort of 642 Japanese patients. **(A)** Archetypes of fat distribution in the Japanese cohort. **(B)** UMAP plot of Japanese patients most similar to each archetype. (C) Number of patients most similar to each archetype or who are intermediate in similarity between archetypes. **(D)** Distributions of fat levels in each depot across archetype groups. **(E)** Distributions of body mass index (BMI) and waist circumference across archetype groups.
